# How the COVID-19 pandemic affected lifestyle and wellbeing factors associated with dementia risk in older adults with Subjective Cognitive Decline and Mild Cognitive Impairment participating in the APPLE-Tree (Active Prevention in People at risk of dementia through Lifestyle, bEhaviour change and Technology to build REsiliEnce) in England

**DOI:** 10.1101/2024.10.28.24316260

**Authors:** Annie Mae Wright, Harriet Demnitz-King, Alexandra Burton, Rachel M. Morse, Sweedal Alberts, Charlotte Kenten, Rosario Isabel Espinoza Jeraldo, Michaela Poppe, Julie Barber, Claudia Cooper

## Abstract

The COVID-19 pandemic negatively affected known dementia risk factors and cognition in older adults. We invited adults with mild cognitive concerns without dementia, aged ≥60 years participating in a randomised controlled trial of a psychosocial, secondary dementia prevention intervention, to complete a co-designed, semi-structured qualitative survey, regarding how the pandemic impacted their lifestyle and wellbeing in areas relevant to dementia risk: social connections, activities, diet, mental and physical health, community and family support. Data was collected between October 2020 and December 2022; we conducted manifest content analysis.

551/746 trial participants completed the survey. Most (n=530, 96%) described pandemic-related changes to lifestyle or wellbeing; two thirds (n=369/545, 67.7%) reported less activities. A quarter (n=145, 26.8%) identified no change in social connections, with others reporting less in-person meetings (n=139, 25.7%) or speaking to less people (n=99; 18.2%); a minority engaged in compensatory online activities (n=31, 5.7%) and online (n=63, 11.6%) or phone (n=90, 16.6%) social contact. Relatively few reported weight gain (n=22, 4.0%); two-thirds reported no change in their diet (n=360, 66.1%), with others eating more unhealthy food (n=31, 4.8%), more food (n=21, 3.9%) and/or snacks (n=11, 2.0%); others reporting more healthy eating (n=26, 4.8%) and/or more home cooking (n=57, 10.5%). Modes of support changed, with reliance on food parcels, online services and shopping by neighbours. Over half reported (almost exclusively negative) mental health pandemic-related changes (n=307, 56.9%), including depression, stress, fear and loneliness; many reported declines in physical health (n=153, 28.1%) and/or fitness (n=70, 12.8%).

Stoical accounts of adaptation and resilience, enabled by technology and community support predominated, but were not possible for all. Reducing the digital divide, tackling loneliness and developing inclusive online/in-person support programmes that are more resilient in future lockdowns could protect cognitively frail people now and in any future pandemic, and contribute to national dementia prevention strategy.

Trial registration: ISRCTN17325135; https://doi.org/10.1186/ISRCTN17325135 (27.11.19)

## Introduction

The COVID-19 pandemic took the lives of over 6.6 million people worldwide. Globally, government measures to limit its spread through social distancing, lockdowns, quarantine and stay at home orders saved lives (1), but the ensuing isolation and anxieties harmed health and wellbeing. People living with dementia were particularly affected; social isolation worsened neuropsychiatric and behavioural symptoms (2) and accelerated functional (2) and cognitive decline (3). Fear of infection, isolation, and closure of dementia services severely affected their wellbeing and that of their family carers (4).

A quarter of the UK population aged 60 and over, live with cognitive impairments that are not dementia but infer an increased risk of dementia (5,6). People meeting criteria for these risk categories are sometimes described as having Mild Cognitive Impairment (MCI), when there are objective cognitive deficits, or Subjective Cognitive Decline (SCD), when deficits are only subjectively measurable. These groups were adversely affected by the pandemic, experiencing increased rates of frailty and depression (7), and reporting more detrimental lifestyle changes, including reduced physical activity, increased smoking and greater alcohol consumption, than people without cognitive impairments (8).

The COVID-19 pandemic resulted in a significant worsening of cognition in older adults, and this was associated with changes in known dementia risk factors (e.g. loneliness, substance misuse, reduced exercise) (9). The National Risk Register emphasises the importance of learning from the pandemic, so that planning for future events is based on a broad understanding of potential health, social, financial and environmental impacts, and community capacity and capabilities to support preparedness, response and recovery, in particular for vulnerable groups (10).

Resilience, defined as the ability to adapt well in the face of difficulties, has been proposed as a defence against loneliness and social isolation during the pandemic, and was probably an important buffer between this and how other pandemic-related stressors affected lives (11). Personal resources (e.g. psychological resilience, self-efficacy) and social resources (e.g. emotional support, social connectedness) can mitigate threats to physical and mental health, social adjustment, and quality of life (11). One study found that participation in physical exercise during the pandemic reduced anxiety in older adults, while social participation supported mental resilience (12). Several studies have reported how technology use, specifically video-calling, buffered pandemic-induced loneliness and isolation (11).

The ability to build resilience is an interaction between individuals and the social environment and should not be construed as an individual achievement (13). Not all older people are equally able to exhibit resilience, leading to new social divisions. For example, those living with mild cognitive concerns may be less able to make changes to reduce dementia risk. An emphasis on agency burdens individuals with the personal responsibility of staying healthy, whether or not this is possible; dementia prevention is a societal concern (14).

In the largest known study on how the pandemic affected the lifestyle and wellbeing of people with mild cognitive concerns to date, we drew on the conceptual framework of resilience (15) to consider how older people experiencing cognitive concerns were able or not to maintain healthy lifestyle behaviours and connections in the dual challenging contexts of cognitive impairments and pandemic-related social restrictions. We recruited participants from a Randomised Controlled Trial (RCT) of a dementia prevention intervention, APPLE-Tree (Active Prevention in People at risk of dementia through Lifestyle, bEhaviour change and Technology to build REsiliEnce) (16). We aimed to explore how older adults with cognitive concerns (i.e. MCI or SCD) were able, or not able to engage in lifestyle activities associated with dementia prevention and maintain their wellbeing. We considered what accounts of resilience (adaptation of routines to pandemic contexts) or challenges to adapting, might tell us about how resilience is best supported and maintained in this population in adverse situations.

## Method

### Study sample

All participants recruited for the APPLE-Tree RCT investigating the effectiveness of a multidomain dementia prevention intervention on reducing cognitive decline in people with cognitive concerns (16) were invited to complete the semi-structured, qualitative survey during the baseline assessment. The APPLE-Tree Trial recruitment took place in participating primary care practices and secondary care memory services; in these settings, which accounted for the majority of recruitment, all eligible participants were approached by letter, targeted at those with some markers of frailty, inviting them to contact researchers if they were worried about their memory, or approached directly by NHS staff (memory services). We also recruited through charities for older people: the Joint Dementia Research Register and social media, newspaper, and online advertisements.

We included people aged 60+, who met criteria for MCI or SCD. This was operationalised as either a Quick MCI test score between 50 and 61 (participants scoring <50 were included if their low scores were consistent with MCI/SCD due to, for example, educational attainment or speaking English as a second language) or, alternatively, a score of ≥62 with a ‘yes’ response to the question, ‘Has your memory deteriorated in the last 5 years? Or has a friend/family member noticed it deteriorating?’ and at least one of the following questions: ‘Is your memory persistently bad, or has a friend/family member noticed it being persistently bad?’ or ‘Are you or others around you concerned about this?’ (17). Further inclusion criteria were a Functional Assessment Questionnaire score <9 indicating no significant cognitive impairment (18) and having a family member/friend/professional to act as an informant who was in contact with the participant at least once a month. An additional inclusion criterion was a willingness to engage in a videocall group intervention for the APPLE-Tree study.

We excluded people with a diagnosis of a primary neurodegenerative disease, advanced, severe unstable or terminal medical condition or severe mental illness, or who lacked capacity to consent. We also excluded people with an AUDIT-C (Alcohol Use Disorders Identification Tool) score ≥8, indicative of harmful alcohol use (19). Lack of access to WIFI or a device to access video calls was not an exclusion criterion as participants were given assistance to access video calls and provided with Mifi and tablets to use throughout the intervention, if required. Full details of the APPLE-Tree trial, including detailed eligibility criteria, are published (27).

### Procedure

Almost all assessments took place by video-call, but when it facilitated participation, a small number were conducted face-to-face where COVID restrictions allowed. We followed government guidance on social distancing during face-to-face interviews (20). Each participant was asked to provide information on their age, gender, ethnicity, and any diagnoses related to memory. As part of the baseline assessment, fully detailed in the main protocol (27), questions were developed in consultation with the APPLE-Tree Patient and Public Involvement group to ask about changes to lifestyle and wellbeing which they attributed to the pandemic. A PPI member piloted the questionnaire. They used as a starting point the lifestyle and wellbeing changes identified as modifiable risk factors for dementia, that were the focus of the APPLE-Tree intervention (21): healthy eating, increased social connections, physical and mental activity, and looking after mental and physical health. Respondents were asked about “recent changes due to COVID-19”; the researcher asked them to compare their current situation to pre-pandemic. The questions asked were: “How did the recent changes due to COVID-19 change:

a. Who you speak to each week?
b. What you eat?
c. What activities you do?
d. How you access help and who you turn to if you need:

- emotional support?
- practical help?
e. Your mental wellbeing (e.g. worries, mood)?
f. Your physical wellbeing?
g. Who you provide care for?”

Responses were free-text; then coded by researchers (see analysis section). Participants could decide to self-complete the questionnaire having been sent this via post or email after meeting with the researcher for the baseline APPLE-Tree interview, or to complete with the researcher, who also recorded notes including verbatim quotes. Thus most quotes in the results section are in participants’ own words, while a small number are in the third person, where the participant opted to reply to questions verbally while the researcher made notes. Unless the participant opted to complete the questionnaire with a friend or family member present, only the researcher and participant were present during interviews. The APPLE-Tree assessment took around 90 minutes, and the COVID questionnaire 5-10 minutes. Interviews were not audio/video recorded, and no additional fieldnotes were taken.

Researchers were from non-clinical, graduate health or sociology backgrounds, most were female, and they were employed at a university to work on the trial and trained in data collection and delivery of the intervention (22). They typically contacted participants before baseline assessments to plan a suitable date or time, but otherwise were not previously known to them. At the start of assessments, they introduced their role, to collect data prior to randomisation to study group, for the purpose of understanding how the pandemic was affecting daily lives. AMW (lead researcher) was a medical student who undertook this study for her dissertation, supervised by CC and RM. RM and SA were study researchers, CC (academic psychiatrist) was Chief Investigator and AB (applied mental health researcher) a co-investigator. CK and HDK were postdoctoral researchers with geography and psychology backgrounds respectively.

### Data analysis

We used manifest content analysis, which involves creating codes directly from the recorded text to analyse the broad surface structure of the data(23). We used Nvivo software to organise data. We generated codes inductively, applying the four stages of content analysis(24): decontextualization, recontextualization, categorisation and compilation. The researcher AMW conducted the analysis and CC independently coded 10% of responses; the level of agreement was >90%. We used the broad categories outlined in our questions (a-g above), inductively coding content within these to develop our coding tree (Appendix 1). In interpreting data, we considered how our findings were situated within our selected conceptual framework of resilience, considering how personal resources, including resilience and social resources influence coping (15).

## Results

### Sample description

Surveys were completed between October 2020 and December 2022. Of 746 APPLE-Tree trial participants, 551 (74%) completed the COVID-19 questionnaire (numbers responding to each question shown in Table 2). Reasons for non-completion were not formally reported, but were typically due to questionnaire fatigue as the survey comprised the final part of the assessment battery. Sociodemographic characteristics of those completing questionnaires were comparable to the baseline trial sample (Table 1). Of the individuals who completed the questionnaire, 132 (24%) did so between October 2020 and July 2021 when a national lockdown was in place, while 419 (76%) participants responded after legal restrictions were removed (August 2021-December 2022) (3). During this second period, there were ongoing social restrictions, for example, face masks were compulsory in public spaces until the end of January 2022. We considered during analysis how timing of the interview might influence data. Most participants (n=512/551, 93%) had access to online video-calling during the pandemic.

**Table 1:**
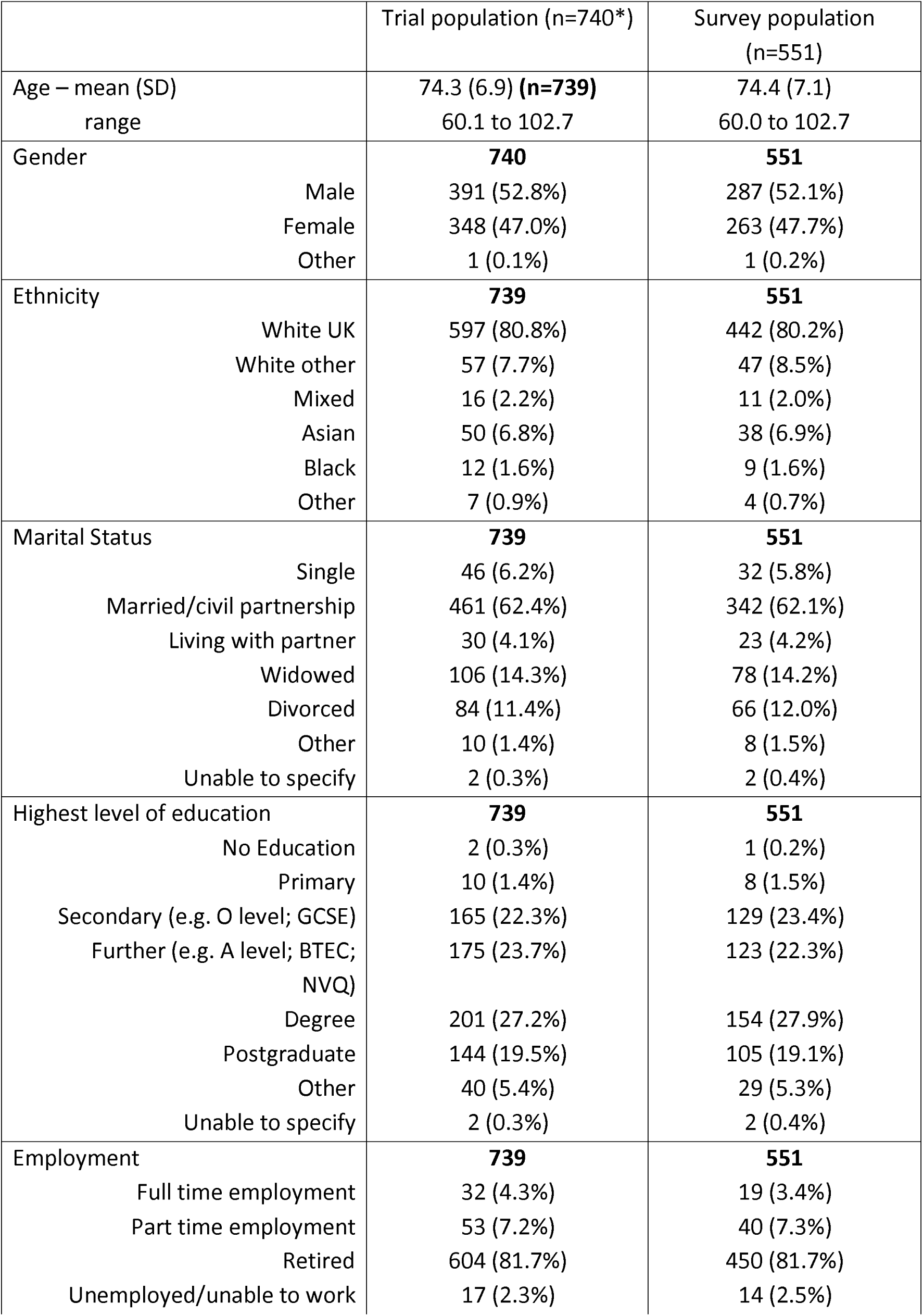
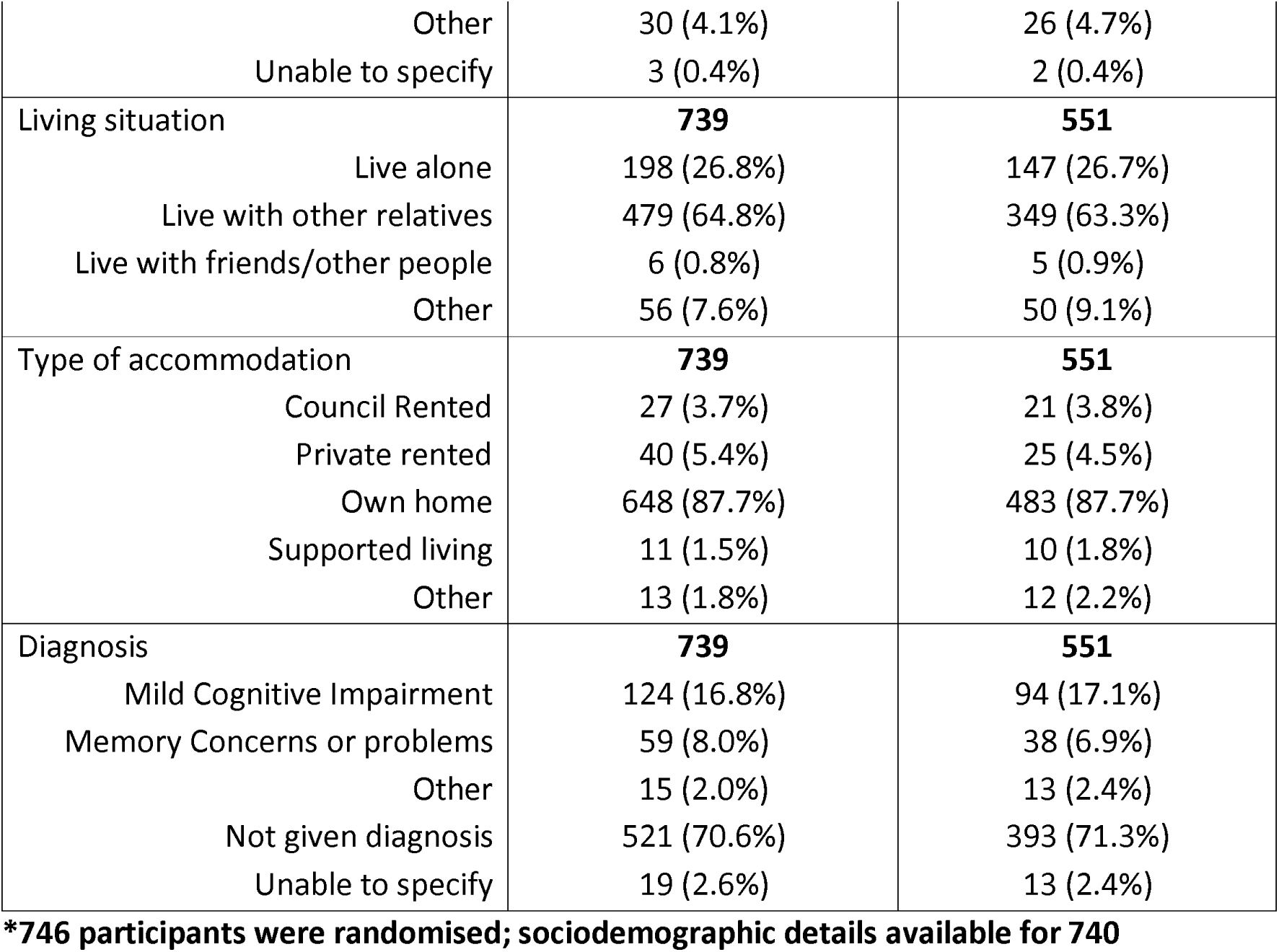
Baseline characteristics of APPLE-Tree trial participants and those participating in the qualitative survey.

|  | Trial population (n=740*) | Survey population (n=551) |
| --- | --- | --- |
| Age – mean (SD)<br>range | 74.3 (6.9) (**n=739**)<br>60.1 to 102.7 | 74.4 (7.1)<br>60.0 to 102.7 |
| Gender | **740** | **551** |
| Male | 391 (52.8%) | 287 (52.1%) |
| Female | 348 (47.0%) | 263 (47.7%) |
| Other | 1 (0.1%) | 1 (0.2%) |
| Ethnicity | **739** | **551** |
| White UK | 597 (80.8%) | 442 (80.2%) |
| White other | 57 (7.7%) | 47 (8.5%) |
| Mixed | 16 (2.2%) | 11 (2.0%) |
| Asian | 50 (6.8%) | 38 (6.9%) |
| Black | 12 (1.6%) | 9 (1.6%) |
| Other | 7 (0.9%) | 4 (0.7%) |
| Marital Status | **739** | **551** |
| Single | 46 (6.2%) | 32 (5.8%) |
| Married/civil partnership | 461 (62.4%) | 342 (62.1%) |
| Living with partner | 30 (4.1%) | 23 (4.2%) |
| Widowed | 106 (14.3%) | 78 (14.2%) |
| Divorced | 84 (11.4%) | 66 (12.0%) |
| Other | 10 (1.4%) | 8 (1.5%) |
| Unable to specify | 2 (0.3%) | 2 (0.4%) |
| Highest level of education | **739** | **551** |
| No Education | 2 (0.3%) | 1 (0.2%) |
| Primary | 10 (1.4%) | 8 (1.5%) |
| Secondary (e.g. O level; GCSE) | 165 (22.3%) | 129 (23.4%) |
| Further (e.g. A level; BTEC; NVQ) | 175 (23.7%) | 123 (22.3%) |
| Degree | 201 (27.2%) | 154 (27.9%) |
| Postgraduate | 144 (19.5%) | 105 (19.1%) |
| Other | 40 (5.4%) | 29 (5.3%) |
| Unable to specify | 2 (0.3%) | 2 (0.4%) |
| Employment | **739** | **551** |
| Full time employment | 32 (4.3%) | 19 (3.4%) |
| Part time employment | 53 (7.2%) | 40 (7.3%) |
| Retired | 604 (81.7%) | 450 (81.7%) |
| Unemployed/unable to work | 17 (2.3%) | 14 (2.5%) |
| Other | 30 (4.1%) | 26 (4.7%) |
| Unable to specify | 3 (0.4%) | 2 (0.4%) |
| **Living situation** | **739** | **551** |
| Live alone | 198 (26.8%) | 147 (26.7%) |
| Live with other relatives | 479 (64.8%) | 349 (63.3%) |
| Live with friends/other people | 6 (0.8%) | 5 (0.9%) |
| Other | 56 (7.6%) | 50 (9.1%) |
| **Type of accommodation** | **739** | **551** |
| Council Rented | 27 (3.7%) | 21 (3.8%) |
| Private rented | 40 (5.4%) | 25 (4.5%) |
| Own home | 648 (87.7%) | 483 (87.7%) |
| Supported living | 11 (1.5%) | 10 (1.8%) |
| Other | 13 (1.8%) | 12 (2.2%) |
| **Diagnosis** | **739** | **551** |
| Mild Cognitive Impairment | 124 (16.8%) | 94 (17.1%) |
| Memory Concerns or problems | 59 (8.0%) | 38 (6.9%) |
| Other | 15 (2.0%) | 13 (2.4%) |
| Not given diagnosis | 521 (70.6%) | 393 (71.3%) |
| Unable to specify | 19 (2.6%) | 13 (2.4%) |

**\*746 participants were randomised; sociodemographic details available for 740**

**Table 2:**
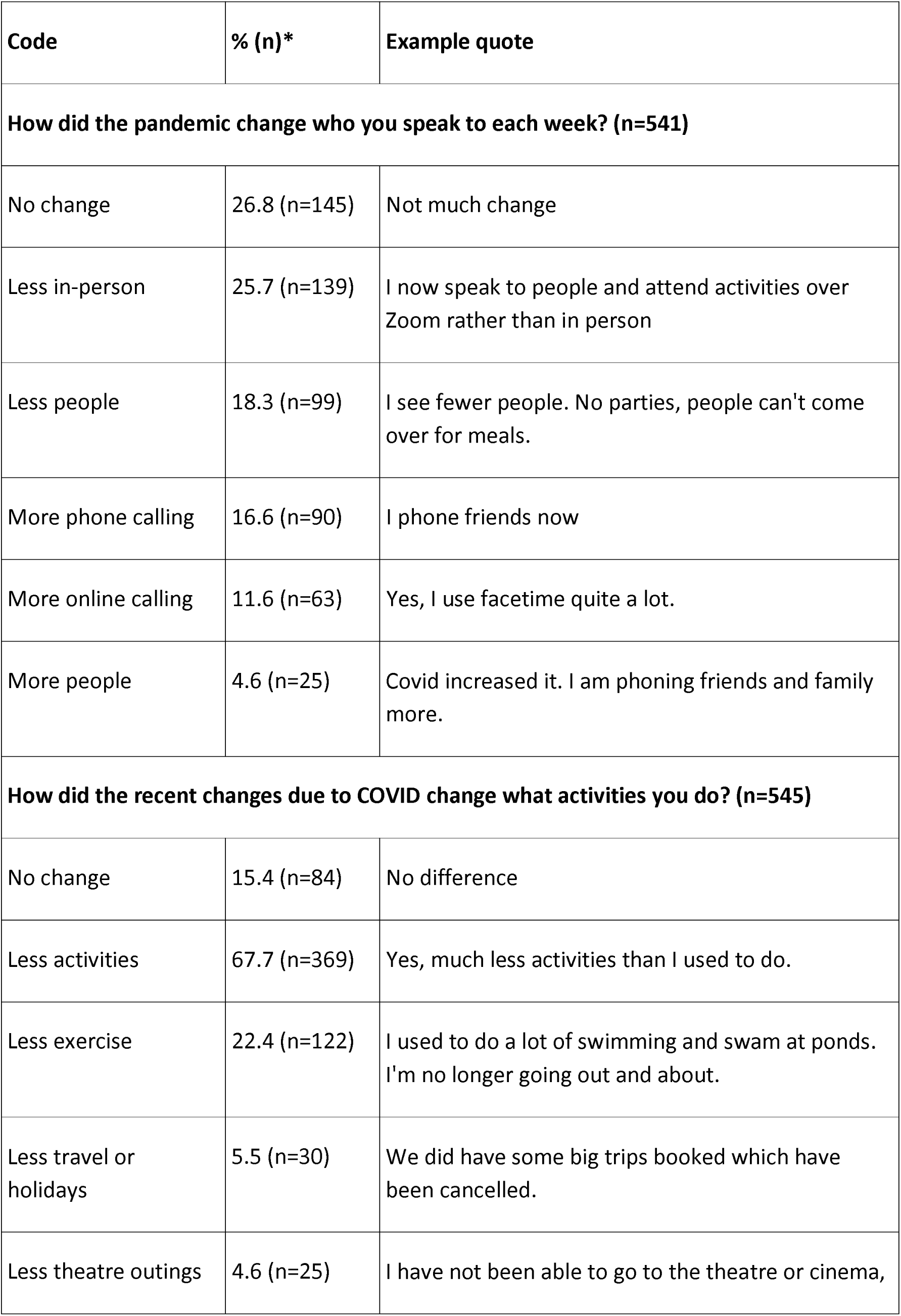
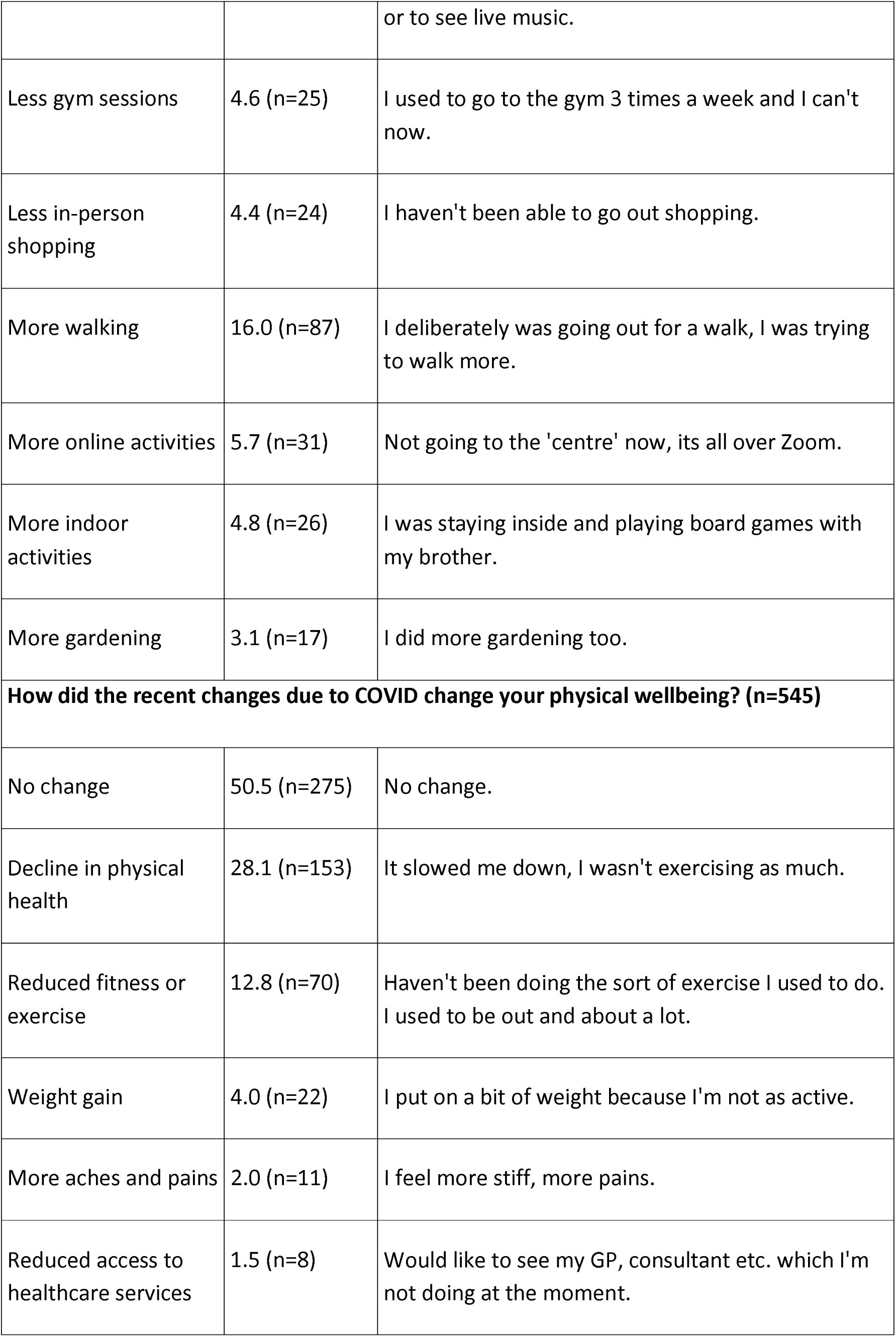
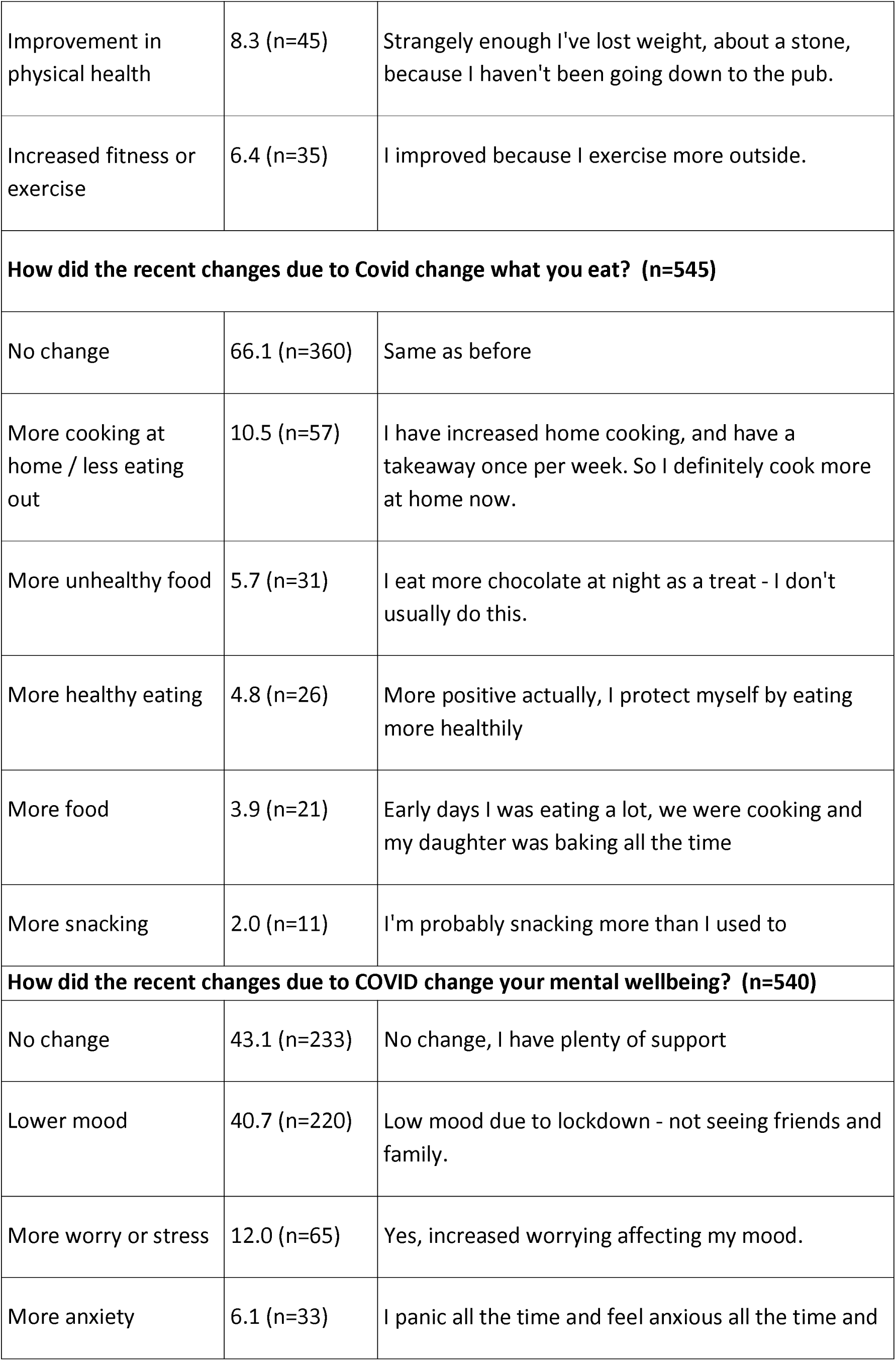
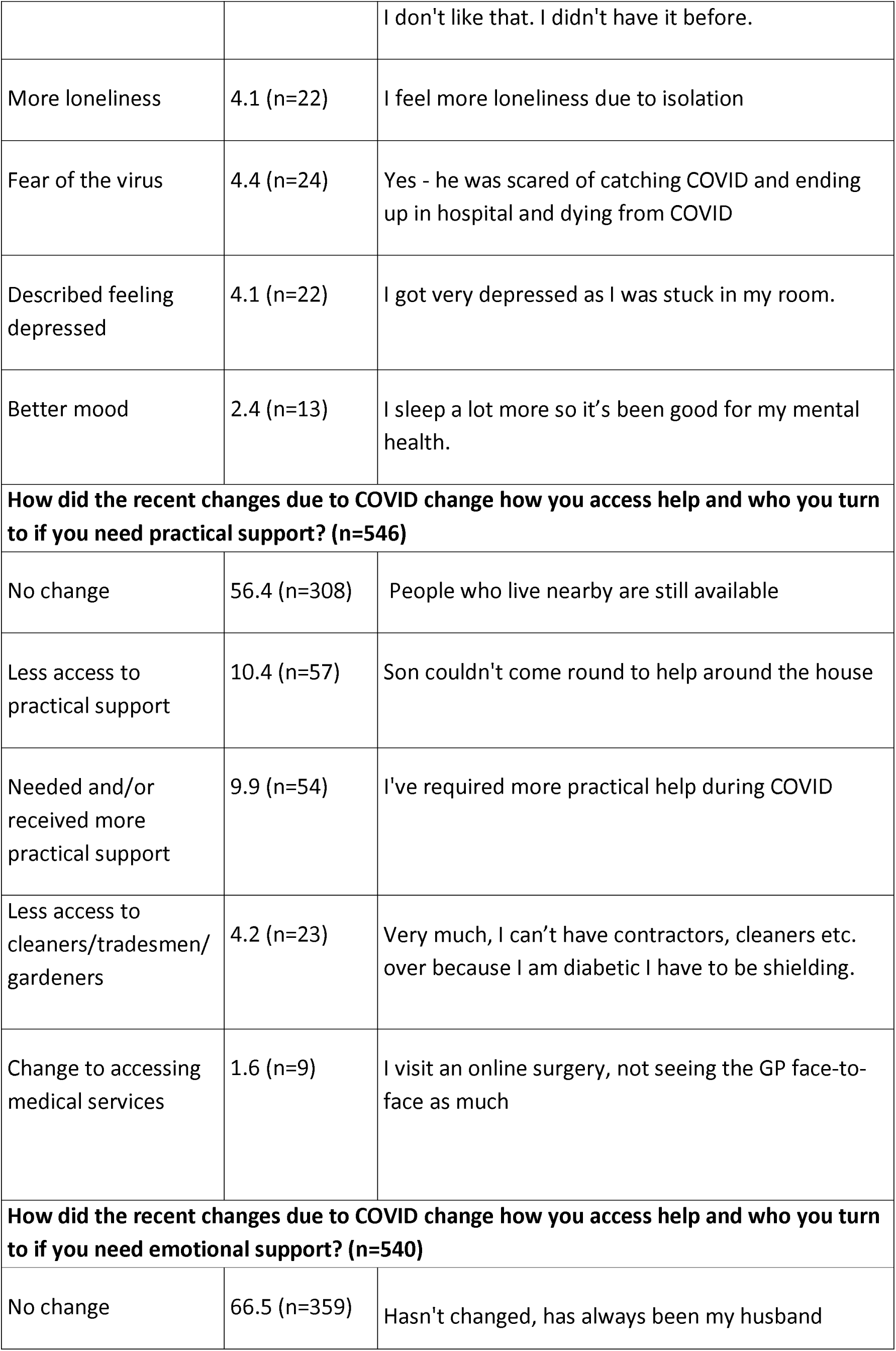
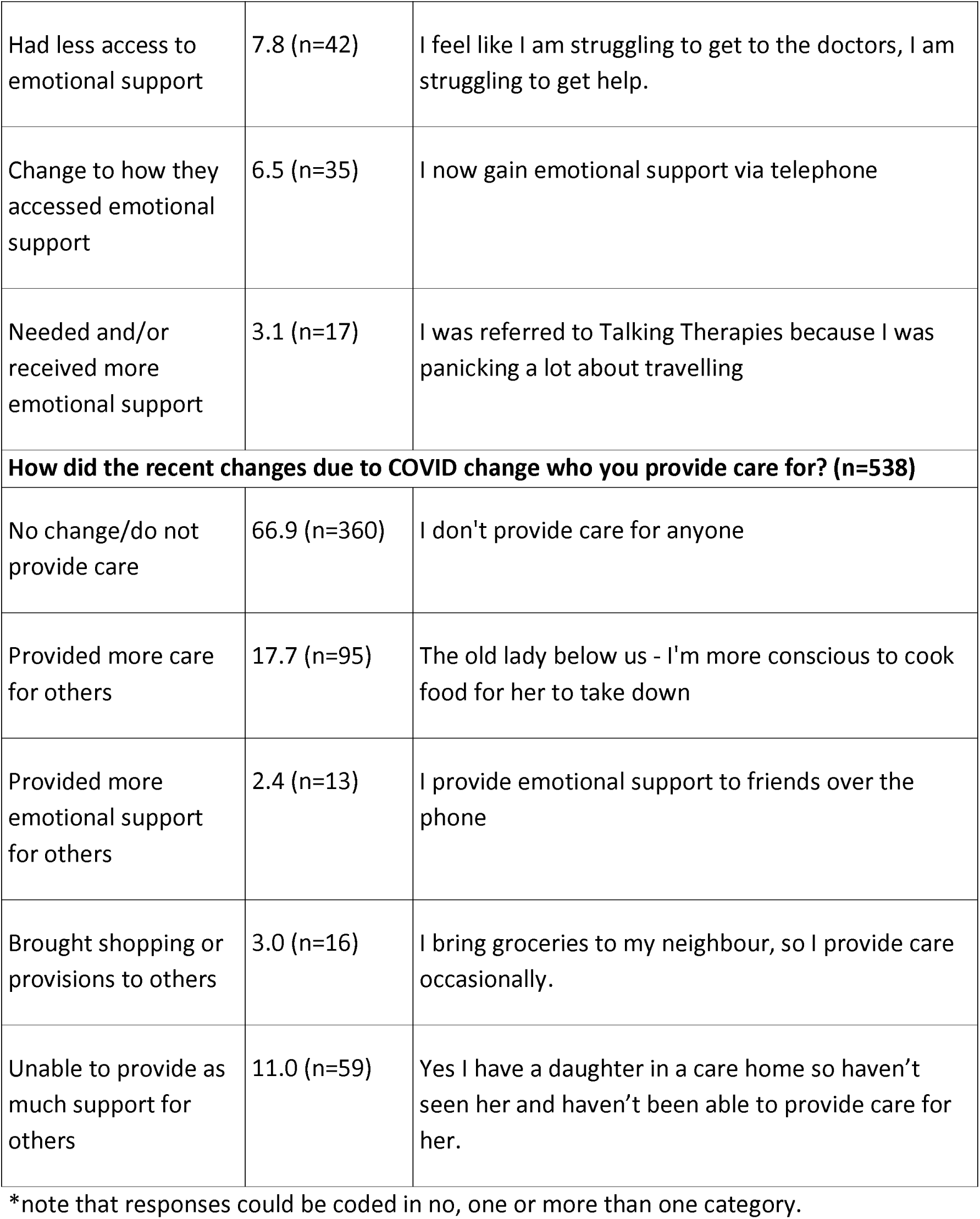
Description of content analysis findings.

### Content analysis findings (Table 2)

530/551 (96%) of participants described experiencing some change to their lifestyle or wellbeing because of the pandemic, while 21 (4%) described no changes; of these, three responses were collected before July 2021 and eighteen after (25).

Content analysis findings are detailed in Table 2, with example quotes, and discussed below. Note that categories reported for each question were not mutually exclusive, respondents could endorse more than one. The coding tree is included in Appendix 1.

#### Changes to who participants spoke to (n=541)

A quarter (n=145, 26.8%) identified no change in social connections, with others reporting less in-person meetings (n=139, 25.7%) or speaking to less people (n=99; 18.3%). The following was a typical response, describing the impact of fewer social contacts:

> *“There was a big change, I speak to less people every week. I am also speaking to less people out and about”* (female, aged 60-64, responded before August 2021).

This was to some extent balanced with more telephone communication (n=90, 16.6%) and more video-call contact (n=63, 11.6%). For a minority, (n=25, 4.6%) the focus on remote (phone or video-call) connections enabled more social connections than pre-pandemic.

> *"I speak often to my mother, brother and sister weekly in a Zoom call, I didn’t do that before” (*male, aged 60-64, responded before August 2021).

Participants who reported no reduction in contact with others during the pandemic often also reported low pre-pandemic levels of contact:

> *“I don’t like speaking to people but I never have”* (female, aged 70-74, responded after July 2021).)

For others, pandemic-related changes of circumstances, such as moving in with a partner, increased contact:

> “*Speak to partner more as staying with him*” (female, aged 75-79, responded before July 2021).)

#### Changes to activities (n=545)

Only 84/545 (15%) of respondents reported that the COVID-19 pandemic restrictions had no impact on their daily activities. For respondents who experienced change, activities outside of their home reduced:

> *“Yea our activities went to nothing, all our clubs and activities closed, even church, we didn’t really do anything”* (female, aged 75-79, responded after July 2021).

Some participants gave additional information as to which activities were affected, citing travel or holidays (n=30, 5.5%), theatre outings (n=25, 4.6%), going to the gym (n=25, 4.6%), and in-person shopping (n=24, 4.4%):

> *“I stopped going to pretty much all local shops, normally I would go two or three times a week and to the supermarket. I still go to the bakers and butchers but I’m restricted in that way”* (male, aged 65-69, responded before July 2021).)

122 (22.4%) reported a reduction in one or more forms of exercise:

> *“Prior to lockdown I was going to the gym 3 times a week, and walking more. I had to shield over lockdown”* (male, aged 75-79, responded after July 2021).

Home based activities increased due to COVID-19 pandemic restrictions. For example, 31 (5.7%) engaged in more online activities; 26 (4.8%) performed more indoor activities and 17 (3.1%) did more gardening. One in five respondents (n=87, 16%) walked more during the pandemic.

> *“I deliberately was going out for a walk, I was trying to walk more”* (female, aged 70-74, responded after July 2021).

These necessary changes to daily activities were often experienced negatively, even when overall activity levels were maintained:

> *“Before this all started, I was making improvements in going out more, felt more alive. I always struggled with anxiety, so this has always been a problem for me. When the pandemic began, I felt much more isolated. I was staying inside and playing board games with my brother”* (male, aged 70-74, responded after July 2021).)

#### Changes to physical wellbeing (n=545)

Just over half of the older adults who answered this question (n=275, 50.5%) reported no change to physical wellbeing; as shown in Table 2, reports of decline (in physical health, in fitness or exercise levels) were prominent in other narratives:

> *“I feel more stiff, more pains”* (female, aged 75-79, responded before July 2021).

8 (1.5%) of participants reported reduced access to healthcare services; this was described by a participant who had:

> *“issues with [their] hands but [it is] hard to see [a] GP about it so [they had] a delayed diagnosis and treatment”* (male, aged 80-84, responded after July 2021).

22 (4%) described weight gain:

> *“I put on a bit of weight because I’m not as active”* (male, aged 65-69, responded after July 2021).

A small number gave responses that we classified as improvements in physical health (n=45, 8.3%), and/or an increase in exercise or fitness (n=35, 6.4%):

> *“Yes – because he was doing his exercises every day and so he was fitter than he had been in a long time”* (male, aged 85-89, responded after July 2021).

The role of socioeconomic factors in enabling resilience was evident in this next response:

> “*We have a large garden, so there was plenty of gardening to do and because I have a workshop, and I am a ‘car nerd’ I usually spend all my time there in the workshop, so luckily covid didn’t upset me because I could still do most of the things I enjoy doing*” (male, aged 80-84, responded after July 2021).)

For another participant, financial stresses made worse by the loss of employment negatively impacted sleep and wellbeing:

> *“Financial worry does keep me awake - like everybody I’m sometimes overwhelmed with the gravity of it” (*female and aged 64.4 years, responded after July 2021)

The loss of many routine medical services impacted physical wellbeing. In this next quote, a participant describes how a regular procedure that prevented a need for a catheter and incontinence was not available for a time in the pandemic:

> “*Doctors were closed down - my botox injections in my bladder stopped and I had a catheter installed, which I did not get on with it. I also went to a&e with my incontinence problems and they could not help either. It was hard*” (male, aged 70-74, responded after July 2021).)

#### Changes to diet (n=545)

A third of respondents (n=360, 66.1%) reported a change in diet.

26 (4.8%) of those that experienced change reported more healthy eating; and/or cooking at home more (n=57, 10.5%):

> *“Before COVID, I used to go out to restaurants, now I cook my own food and I think I eat healthier, more fruits and veggies”* (male, aged 60-64, responded after July 2021).

31 (5.7%) ate more unhealthy foods including more chocolate (n=8), cake (n=8), biscuits (n=4), and processed food (n=5). Participants discussed these unhealthier eating habits:

> *“Never used to snack, now snacks more out of boredom/ being indoors”* (female, aged 65-69, responded before July 2021).
>
> *“I ate more sweets and chocolate than I should have”* (female, aged 75-79, responded after July 2021).

21 (3.9%) increased their overall food intake and 11 (2.0%) snacked more. One respondent described eating less, linking this to a loss of routine and confusion. Their response indicated a potential role of cognitive impairment as a barrier to the resilience evident in other responses, where routines were adapted, for example in this next quote:

> *“I think I eat less, I don’t know what to eat sometimes, I feel confusion around eating because I am home all day” (female, aged 70-74, responded after July 2021).)*

One respondent explained how they managed to maintain weight loss despite not having access to their support group:

> *“I was really good during lockdown, I was in the Slimming World group but I couldn’t go, I managed to stay the same, I was stable on my weight, I didn’t lose but I kept it under control” (female, 60-64, responded after July 2021).)*

#### Changes to mental wellbeing (n=540)

Nearly half of participants (48%, n=257) described (almost exclusively negative) changes in their mental health due to the pandemic. The most frequently reported issues were lower mood (n=220, 40.7%), increased worry or stress (n=65, 12%), heightened anxiety (n=33, 6.1%), fear of the virus (n=24, 4.4%), and depression (n=22, 4.1%). The decline in mental wellbeing can be seen in the following response:

> *“Since COVID started I’ve been put on antidepressants and the dosage has increased during the last 6 months. It’s the stress of COVID … My anxiety and depression has got a lot worse during COVID”* (female, aged 60-64, responded before July 2021).

22 (4.1%) of participants mentioned feeling more lonely when asked about their mental health. This next quote indicates how anxiety and isolation could be mutually reinforcing, creating a vicious cycle:

> *“I feel lonely, I’m scared, it’s changed a lot - when you sit in a room and can’t go out because you think you might catch it, I feel down, mentally keep thinking I won’t live any longer”* (male, aged 75-79, responded before July 2021).

Only 13 (2.4%) who experienced change reported improved mental health during the pandemic:

> *“I think on the whole I felt happier as I had more time to myself and time to think”* (male, aged 75-79, responded after July 2021).

#### Changes to practical support (n=546)

Just over half (56.4%, n=308) of participants reported experiencing no change in how much they accessed practical support. Other responses described having less access to support from tradespeople and different means of accessing medical services (Table 2).

> *“[I] haven’t called in many professionals (gas service, cleaner etc.) so reduced the practical help that I would have wanted like repairs”* (male, aged 65-69, responded before July 2021).

For those who reported receiving more practical support, this was from family and friends who, for example, delivered shopping or meals.

> *“I’ve required more practical help during COVID e.g. my friends and family help me with shopping now as I’m classed as vulnerable”* (female, aged 60-64, responded before July 2021).

Many participants mentioned using more online shopping, an option that would not have been available to those on lower incomes. For others, there was a need to rely on “food parcels” (male, aged 60-64, responded after July 2021), or friends and family. Some commented on the frustrations of this necessary reliance and increased dependency:

> *“Yes, had to rely on daughter bring in me food, so whatever she would get from the supermarket and I prefer to do it myself”* (female, aged 80-84, responded after July 2021).)

#### Changes to emotional support (n=540)

A fifth of participants (22%; n=119) experienced a change in how much emotional support they received; other respondents (n=42) indicated they had less access to emotional support during the pandemic.

One participant, whose response was recorded verbatim by the researcher in an interview completed by video-call, described greater support needs but receiving less support:

> *“She felt really desperate and that’s why she called the IAPT [Improving Access to Psychological Therapies] service but no response. She didn’t get any help. She feels horrible and angry most of the time”* (female, aged 65-69, responded before July 2021).

Others (n=35) reported a change to how they accessed emotional support with less face-to-face support. For example, one participant reported continuing support from: *“The GP, but the way I was seeing the GP changed (virtually)” (male, aged 70-74, responded after July 2021).)*

Remote connections did not always compensate for in-person contact. For some, changes in GP accessibility felt less supportive:

> *“I feel like I am struggling to get to the doctors, I am struggling to get help. It feels like doctors care less, just prescribing”* (female, aged 70-74, responded after July 2021).)

Several participants described missing physical contact:

> “*I’m not able to hug my children and grandchildren*” (male, aged 75-79, responded before July 2021).)

Some (n=17) participants reported needing and receiving more emotional support; emotional and practical support was often provided by neighbours:

> *“I’ve discovered how brilliant my neighbours are, they’re 6 different neighbours and they check up on me and bring me cooked food”* (female, aged 70-74, responded before July 2021).)

#### Changes to provision of support to others (n=538)

Around two-thirds 66.9% (n=360) of respondents felt that COVID had not changed the support they provided to others; others (n=95) provided more care, and/or more emotional support for friends and family (n=13) or brought shopping or provisions to others (n=16):

> *“The old lady below us – I’m more conscious to cook food for her to take down”* (female, aged 65-69, responded before July 2021).

Others (n=59) were unable to provide as much support to others as they used to, mainly because they were unable to have contact with grandchildren or children they usually provided care for:

> *“Yes I have a daughter in a care home so haven’t seen her and haven’t been able to provide care for her”* (female, aged 65-69, responded before July 2021).

## Discussion

This is, to our knowledge, the largest survey asking older people living with mild cognitive concerns how government imposed COVID-19-related restrictions affected their lifestyle and wellbeing. Unsurprisingly, most respondents saw fewer people and were less active. Many participants adapted their daily routines, substituting activities that were no longer possible in the pandemic with home cooking, walking, gardening, or online activities. Reports of reduced physical wellbeing, attributed to lower activity and exercise levels, and reduced mental wellbeing (e.g. low mood and anxiety), related to anxieties around the virus and loneliness were common.

Between the stressor (pandemic lockdowns and fears) and accounts of adaptation and accommodating to new contexts, lies “the resilience itself” (15). Social, digital, and community capital facilitated resilience. Most participants did not report any changes to the quantity of practical and emotional support they received, though systems for obtaining these changed. Some respondents compensated for less in-person contact by adopting video-call technology and using the phone more to connect with friends, families, and health professionals. A minority spoke of the distress of isolation, loss of opportunities to connect outside the home, and reduced access to routine health care, indicating that while stoical accounts of adaptation and resilience predominated, this was not always possible.

One account indicated that greater cognitive impairment (confusion around new routines) may have impeded resilience. Social and digital exclusion affect those experiencing socioeconomic deprivation more, as financial resources facilitate access to transport, digital connectivity, health, community and local social services (32). While we did not measure socioeconomic barriers to resilience, or indeed resilience directly, they were suggested by some responses. Examples include a quote above where having a garden and a shed for hobbies supported resilience, and another where financial insecurity was a source of worry and sleeplessness. Online shopping helped many, while for others (who may have not had access to this for reasons including limited financial resources), greater reliance on friends, family, or food parcels from community organisations brought sometimes unwelcome reliance on others. Concerns about physical deterioration due to reduced availability of routine care reflect reports of people living with long term conditions during the pandemic (30).

Though older people with mild cognitive concerns are often highly motivated to reduce dementia risk, they are less likely to be able to do so successfully without support (31). One respondent described how she had managed to maintain, but not progress her goal for weight reduction without the support of her slimming group, which closed in the pandemic. Her account illustrated the importance of social support in achieving lifestyle change. By increasing reliance on remote connectivity, the pandemic reinforced effects of digital exclusion. The 2025 UK digital switchover will eliminate landlines, potential lifelines for many older people in the last pandemic (33), so this will be a pertinent issue in future preparedness.

### Public health and policy implications

Global pandemics such as COVID-19 are relatively rare, but research has predicted an increased frequency in future (34). Despite UK government investment in infrastructure and economic growth to aid post-pandemic recovery, there has been little discussion of strategies to address pandemic-related health repercussions (46). The UK COVID-19 Public Inquiry Module 1, covering Resilience and Preparedness, concluded that Emergency planning failed to sufficiently consider health and social inequalities, and local authorities and volunteers were not adequately engaged (35).

The ongoing Darzi independent investigation into the state of the NHS, which aims to inform a new NHS ten year plan (36) is an opportunity to focus on the prevention of dementia. This needs to include population-wide, primary prevention as well as public health messaging and interventions targeting at risk groups, including those with memory concerns.

While the particular challenges to people living with dementia were acknowledged in the pandemic, the specific needs of people with mild cognitive impairments were not specifically acknowledged (37). Levels of psychological resilience and emotional wellbeing of people with mild cognitive concerns appear to be greater (26–28) than for people living with dementia (4); but lower than for older people without cognitive concerns who reported good social support and drawing on previous coping strategies and life experience throughout the first wave of the pandemic in a previous study (29).

Future preparedness strategies should actively consider those living with mild cognitive concerns, who are often excluded from health and care planning; but whose resilience can be enhanced by community and digital connectivity. Ensuring that the population can be resilient to change may mean integrating online options (and providing skills training where needed) and in-person activities so any future shifts in the form of engagement are less disruptive to the routines of vulnerable groups. This would also be a short-term strategy to reduce dementia risk.

## Limitations

Researchers summarised participants’ responses, which may have introduced bias. However, using the manifest analysis method, responses were coded based on what was said rather than interpreted. Although the survey asked specifically about recent changes due to COVID, level of restrictions varied over the study period, so respondents would have been experiencing these differently depending on when they completed it.

Respondents are unlikely to be a representative sample of the older population living with memory loss, as trial populations are not representative (48). A criterion for participation in the APPLE-Tree trial was willingness to engage in a video-call intervention. Devices were available for loan, but nonetheless those with their own devices, accustomed to online communication, were probably more likely to take part, as were those more amenable to group participation. A telephone interview study of older adults with MCI or SCD in Italy, reported fewer ongoing social activities, perhaps reflecting a greater proportion of people who were digitally excluded in their sample population, or differences in the level of pandemic social restrictions between the UK and Italy (26). Despite these caveats, likely to have biased our sample towards greater resilience (through more socioeconomic, technology, and community capital), we believe our findings can help inform strategies for secondary dementia prevention, including preparedness for future global disasters. While we consider how technology and social connections increased resilience, and discuss evidence that these barriers are often socioeconomically determined, we did not directly compare reports of lifestyle changes against socioeconomic characteristics in this descriptive study.

## Conclusion

We describe the accounts of people living with mild memory loss regarding how their wellbeing and lifestyle were influenced by the COVID pandemic. These evidenced increased loneliness, isolation, and physical and mental distress, but also resilience, through moving to online social connections and adapting new daily activities. Our findings may inform how we protect people with mild cognitive concerns in future disasters. Reducing the digital divide and tackling loneliness in older people now could reap dividends, including in any future pandemic.

## Supporting information

Coding Tree

## Acknowledgements

We would particularly like to thank the APPLE-Tree community of interest and the coproduction group.

## Statements and Declarations

### Declaration of Conflicting Interests

The author(s) declared no potential conflicts of interest with respect to the research, authorship, and/or publication of this article.

### Funding

This work is funded by an Economic and Social Research Council/National Institute for Health Research programme grant (ES/S010408/1).

### Ethical approval and informed consent statements

We obtained approval from the London Research Ethics Committee (Reference 19/LO/0260) and the UK Health Research Authority and written or audio-recorded consent from all participants.

### Data availability statement

Data is available from the corresponding author on receipt of a reasonable request.

## Notes

### Competing Interest Statement

The authors have declared no competing interest.

