## Supplementary material for "How the COVID-19 pandemic affected lifestyle and wellbeing factors associated with dementia risk in older adults with Subjective Cognitive Decline and Mild Cognitive Impairment participating in the APPLE-Tree (Active Prevention in People at risk of dementia through Lifestyle, bEhaviour change and Te": Coding Tree

**Appendix 1: Coding Tree**

Changes in who you speak to each week

| Key |
| --- |
| N = No change |
| = No answer / didn't answer question |
| SP = Same people |
| Online = More online calling |
| Phone = More phone calling |
| LI = Less in-person |
| LP = Less people |
| MP = More people |
| L/M = Some people more, some people less |
| LO = Less often |
| MO = More often |
| I = Feel more isolated |

Changes in what you eat

| **Key** |  |
| --- | --- |
| N | no change |
|  | no answer / didn't answer question |
| HE | healthy eating |
| CO | cooked at home more |
| RE | ate out less |
| MF | more food |
| MS | more snacks |
| UN | more unhealthy food |
| CH | chocolate |
| CA | cake |
| RM | ready meals/processed food |
| BI | biscuits |
| C | other change |

Changes in what activities you do

| **Key** |  |
| --- | --- |
| N | no change |
|  | no answer / didn't answer question |
| LA | reduction in one or more activities |
| LS | less socialising |
| LE | less exercise |
| G | no gym |
| TH | no theatre |
| L | no library |
| CC | no community centre |
| S | no shopping |
| M | no music/concerts |
| F | no football matches |
| TR | no travelling/holidays |
| O | more online activities |
| IA | more indoor activities |
| ME | more exercsie |
| MW | more walking |
| R | found alternative exercise |
| MG | more gardening |

Practical help

| **Key** |  |
| --- | --- |
| N | no change |
|  | no answer / didn't answer question |
| LS | less access to practical help |
| MS | had/needed more support |
| D | didn't need practical help |
| SS | had help with shopping/food |
| NC | restricted access to cleaners/tradesmen/gardeners |
| GP | difficulty accessing medical services |
| S | had support from family/friends |
| EX | had help from external services |

Emotional support

| **Key** |  |
| --- | --- |
| N | no change |
|  | no answer / didn't answer question |
| LS | less access to EMOTIONAL? help |
| MS | had/needed more support |
| D | didn't need practical help |
| SS | had help with shopping/food |
| NC | restricted access to cleaners/tradesmen/gardeners |
| GP | difficulty accessing medical services |
| S | had support from family/friends |
| EX | had help from external services |

Mental wellbeing

| **Key** |  |
| --- | --- |
| N | no change |
|  | no answer / didn't answer question |
| ? | some change, don't specify type of change |
| WM | worse mood |
| BM | better mood |
| W | more worried/stressed |
| F | fear of COVID |
| A | more anxious |
| FR | frustrated |
| AN | angry |
| B | more bored |
| D | more depressed |
| L | isolated/lonely |
| I | more irritable/moody |
| WS | worse sleep |

Physical wellbeing

| **Key** |  |
| --- | --- |
| N | no change |
|  | no answer / didn't answer question |
| ? | some change, don't specify type of change |
| BP | better physical health |
| WP | worse physical health |
| A | increased aches/pains |
| MS | reduced access to medical services |
| WG | weight gain |
| ME | more exercise |
| RE | reduced exercise |

Providing support

| **Key** |  |
| --- | --- |
| N | no change |
|  | no answer / didn't answer question |
| P | providing more care |
| L | unable to provide as much care |
| D | don't care for anyone |
| E | providing more emotional support |
| S | bringing provisions to others |
| B | cannot provide childcare |
